# High heritability of ascending aortic diameter and multi-ethnic prediction of thoracic aortic disease

**DOI:** 10.1101/2020.05.29.20102335

**Authors:** Catherine Tcheandjieu, Ke Xiao, Helio Tejeda, Julie A. Lynch, Sanni Ruotsalainen, Tiffany Bellomo, Madhuri Palnati, Renae Judy, Derek Klarin, Rachel Kember, Shefali Verma, Regeneron Genetics Center, Aarno Palotie, VA Million Veterans Program, FinnGen Project, Marylyn Ritchie, Daniel J. Rader, Manuel A. Rivas, Themistocles Assimes, Philip Tsao, Scott Damrauer, James R. Priest

## Abstract

Enlargement of the aorta is an important risk factor for aortic aneurysm and dissection, a leading cause of morbidity in the developed world. While Mendelian genetics account for a portion of thoracic aortic disease, the contribution of common variation is not known. Using standard techniques in computer vision, we performed automated extraction of Ascending Aortic Diameter (AsAoD) from cardiac MRI of 36,021 individuals from the UK Biobank. A multi-ethnic genome wide association study and trans-ethnic meta-analysis identified 99 lead variants across 71 loci including genes related to cardiovascular development (*HAND2, TBX20)* and Mendelian forms of thoracic aortic disease (*ELN, FBN1)*. A polygenic risk score predicted prevalent risk of thoracic aortic aneurysm within the UK Biobank (OR 1.50 per standard deviation (SD) polygenic risk score (PRS), p=6.30×10^−03^) which was validated across three additional biobanks including FinnGen, the Penn Medicine Biobank, and the Million Veterans Program (MVP) in individuals of European descent (OR 1.37 [1.31 - 1.43] per SD PRS), individuals of Hispanic descent (OR 1.40 [1.16 - 1.69] per SD PRS, p=5.6×10^−04^), and individuals of African American descent (OR 1.08 [1.00 - 1.18] per SD PRS, p=0.05). Within individuals of European descent who carried a diagnosis of thoracic aneurysm, the PRS was specifically predictive of the need for surgical intervention (OR 1.57 [1.15 - 2.15] per SD PRS, p=4.45×10^−03^). Using Mendelian Randomization our data highlight the primary causal role of blood pressure in reducing dilation of the thoracic aorta. Overall our findings link normal anatomic variation to extremes observed in Mendelian syndromes and provide a roadmap for the use of genetic determinants of human anatomy in both understanding cardiovascular development while simultaneously improving prediction and prevention of human disease.

## INTRODUCTION

The thoracic aorta originates from multiple developmental anlage to become the largest vascular structure in the human body which receives and distributes oxygenated blood pumped by the left ventricle^1^. Body surface area is a primary determinant of thoracic aortic size, which increases in diameter with the physiological demands that accompany normal growth throughout childhood and into adult life^2^. Extremes in thoracic aortic diameter can manifest in clinically significant disease ranging from thoracic aortic dilation or aneurysm which may be asymptomatic until the occurrence of rupture or dissection^3–5^.

Common genetic variation has been associated with echocardiographic measurement of aortic root diameter within European populations^6^. Heritable differences in the size of the aortic root are present within family members of individuals with congenital malformations of the aorta^7^, and clinical genetic testing for known mendelian forms of aortopathy are an integral part of multidisciplinary care for thoracic aortic disease^8^. However, for up to 75% of individuals affected with thoracic aortic disease, pathogenic mendelian variation is not found on clinical genetic testing and risk factors such smoking and hypertension are often insufficient to explain incident pathology^9^ which suggests that a substantial portion of risk for thoracic aortic disease has yet to be described.

Here we report a genome wide association study of ascending aorta diameter (AsAoD) – extracted using machine learning – from a population-based study of 36,021 cardiac MRI prospectively obtained by the UK Biobank organization. We report 71 new genomic risk loci associated with ascending aorta diameter which explain a large proportion of the total heritability in observed variation in the AsAoD and showed that the polygenic risk score of the AsAoD is predictive for the risk of thoracic aortic aneurysm across multiple populations.

## RESULTS

### Ascending Aorta Diameter (AsAoD) measurement

The AsAoD was derived from a transverse MRI image of the thorax (figure 1a) using a simple segmentation-based approach (see methods section) in 36,021 UK Biobank participants. Overall, in concordance with existing normal data of aortic size, the mean of the AsAoD was 30.6 mm (sd=3.9) in females and 33.3 mm (sd=4.2) in males (figure 1b-1c).

**Figure 1:**
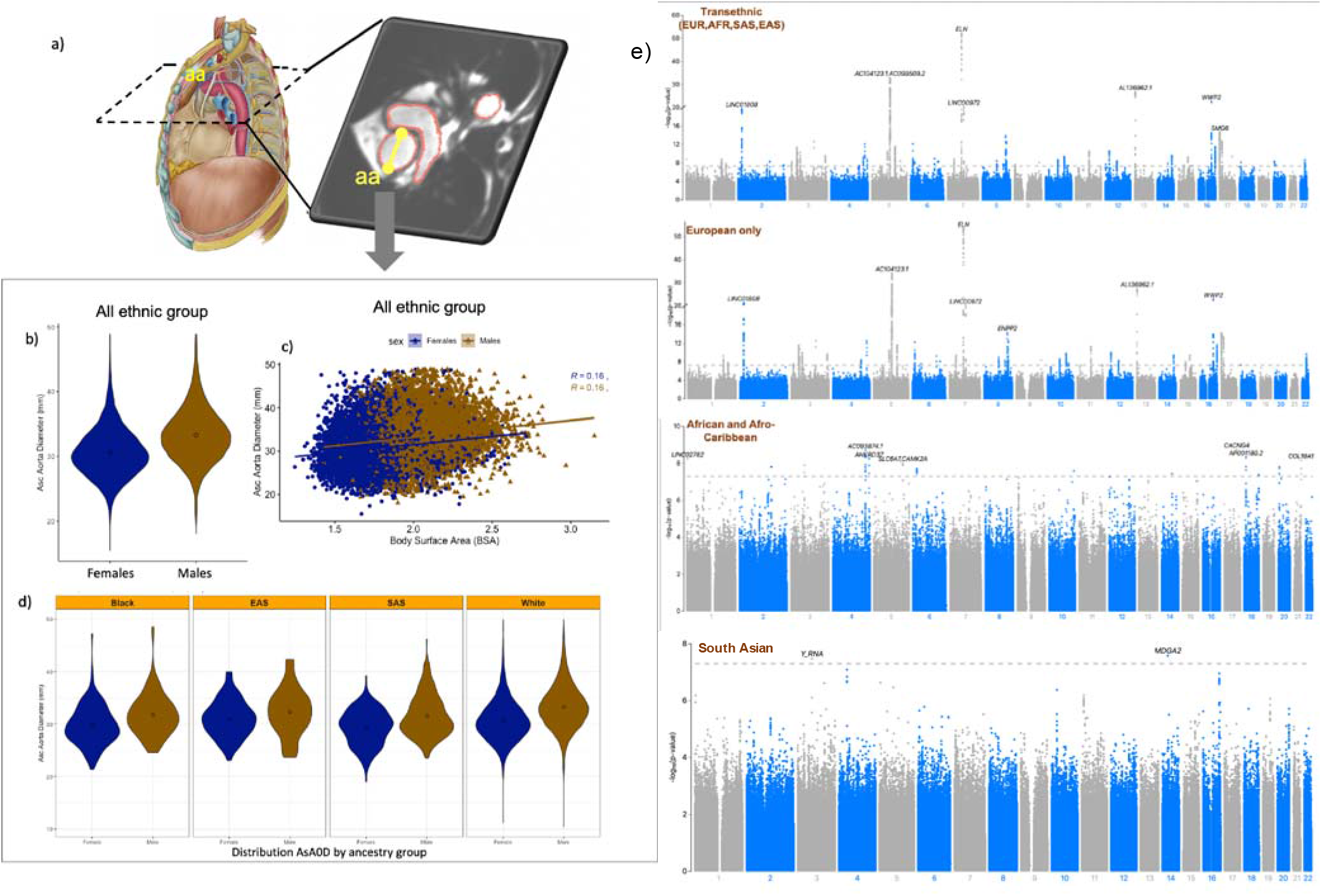
(a) Transverse plane image of the thorax from cardiac MRI, label in yellow is ascending aorta diameter (aa). (b) Violin plots ascending aorta diameter area in female (red) and in male (blue) (table S1). (c) Scatter plot of ascending aortic diameter by body surface area in female (red) and in male (blue). As expected from the general population^10,11^, females had on average a smaller aorta area than males. (d) distribution of AsAoD by genetic ancestry (e) Manhattan plots of AsAoD ethnic-specific GWAS and the trans-ethnic meta-analysis. The dashed line represents a standard threshold of significance (5×10^−08^)

### Ancestry-specific and trans-ethnic meta-analysis of AsAoD

We first performed a genome wide association study (GWAS) of AsAoD adjusted for body surface area in unrelated white European (n = 32,215) in the UK Biobank observing a striking set of 163 independent variants across 42 genomic loci (figure 1e, figure S1, table S2). Using GREML-LDMS within the Europeans, we estimated a high-level of heritability at 0.398 (se=0.072) derived from individual level imputed and genotyped variants with a contribution of variants spread across all range of allele frequencies and a strong contribution of common variants (MAF>0.1) in the heritability estimate (figure S1). Therefore, we elected to perform a GWAS of AsAoD in the African and Afro-Caribbean (n = 262), East Asian (n = 133) and South Asian participants (n = 441) with MRI data in the UK biobank followed by a transethnic meta-analysis of all individuals (figure S2).

We identified a total of 71 genomic risk (table 1) loci associated with AsAoD among which 45 signals were consistent across all ethnic groups (p_het_ > 1e-03), three of these loci reach genome wide significance in the trans-ethnic meta-analysis, 37 loci were significant in the European GWAS; and six loci with African specific lead variants had candidate variants in or near the locus in another ancestry group (table 1, figure S2, table S2). Five genomic risk loci were present only in Europeans while 18 loci were African/Afro-Caribbean specific risk loci among which 3 loci (5q35 (*CAMK2A*), *18q11*(*NAPG*), and 21q22 (*COL18A1*)) were suggestive in Europeans with consistent directionality across all ethnic strata but with significant heterogeneity in the transethnic meta-analysis (figure S3, table S2). Two loci were significant in South Asians with the direction of effect consistent with other ethnic strata but displaying high heterogeneity in the trans-ethnic meta-analysis.

The strongest signal was found in the locus 7q11.2 near Elastin (*ELN*), a gene responsible for elastin production, a key component of elastic fiber structure of vascular tissue including the aorta. The second strongest independent signal was observed in the locus 5p23 near *PCSK1* a proprotein Convertase Subtilisin/Kexin Type 1. Within the African/Afro-Caribbean strata the two strongest signals were found in the locus 4q32 (*AGAP1*) and 4q35 (*ANKRD37*) overlapping the 4q deletion syndrome region which includes a key transcription factor in cardiac development HAND2^12–14^. Interestingly, we observed a separate distinct GWAS signal in Europeans at the 4q32 locus (table 1, figure S3) with effect sizes consistent across all ancestries (table S3).

**Table 1:**
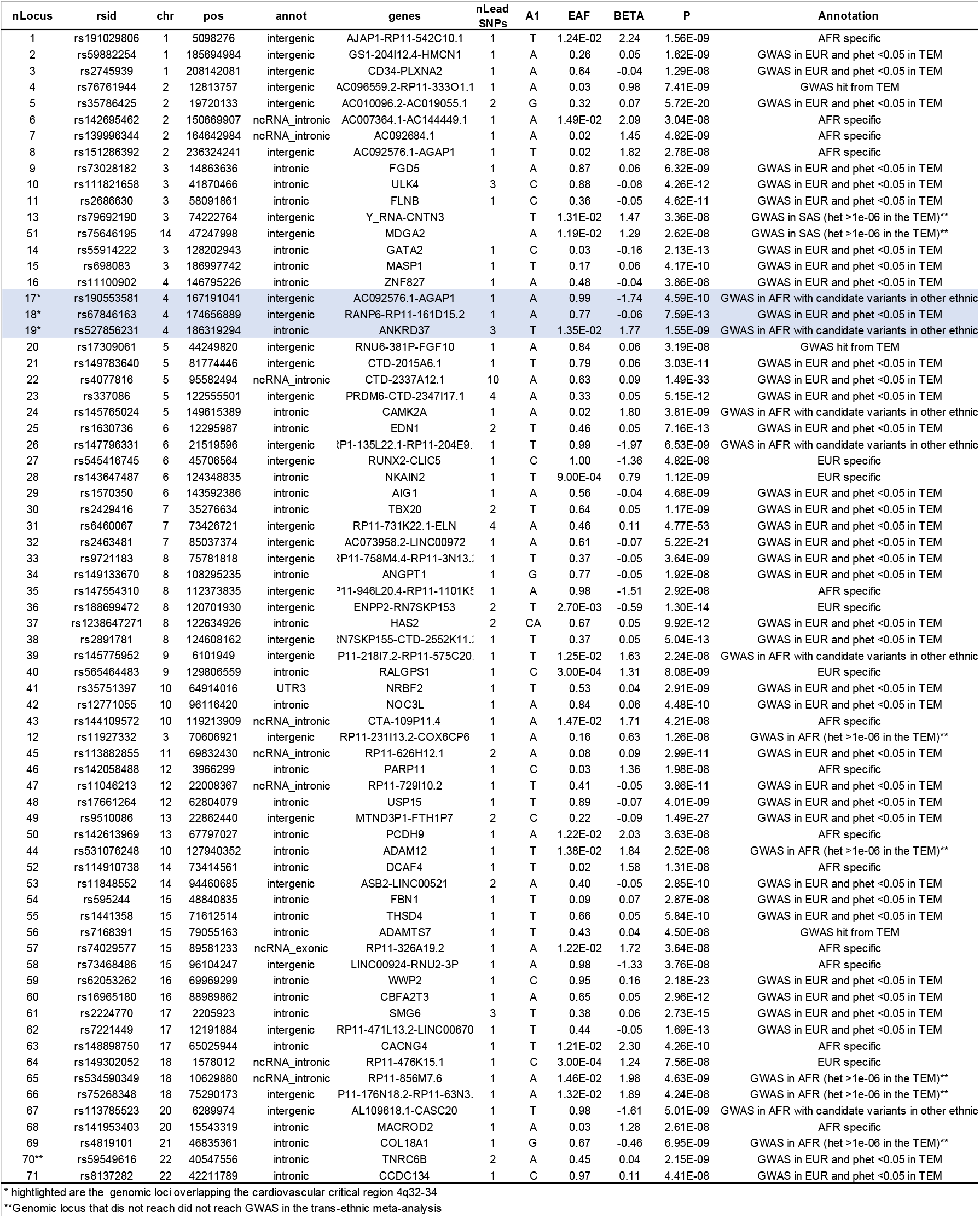
Lead variants in each genomic loci for ancestry specific and the trans-ethnic meta-analysis of AsAoD.

### Variant function prediction and chromatin interaction

To identify variants which alter gene expression or protein function we annotated variants with p<1×10^−05^ using Haploreg^15^. Overall, we identified 16 non-synonymous variants located in several genes including *ELN, ULK4*, and *SMG6*; six variants in promoter regions of *MASP1, TBX20, CDKN1A*, and *HAS-AS1*. Several other variants were located in transcription factors binding sites (TFBs) or in region with high chromatin interaction targeting promoter or enhancer of several genes (table S4).

To further investigate the relationship between genes inside or outside the genomic risk loci, in each independent signal we mapped genes whose promoter or enhancer overlapped with independent variants displaying significant eQTL or chromatin interactions using FUMA^16,17^. Chromatin interaction and eQTLs were observed for multiple loci (figure S4). For instance, strong chromatin interactions were observed between enhancer and promoter of *PCSK1, ELL2, ERAP1, ERAP2, RHOBTB3* and *GLRX* in the genomic risk locus 5p23. Similarly, chromatin interaction was observed between *ELN* in the locus 7q11 and enhancers or promoters of several genes including *TBL2, LIMK2, CLIP2*. Additionally, variants in *ELN, LAT2* and *GTF2IRD1* display significant eQTLs in multiple tissues including aorta (figure S4).

### Genes enrichment and transcriptome-wide association analysis

We used MAGMA to leverage our summary statistics to examine gene-level associations, testing 19,288 annotated genes tagged by variants from our GWAS summary. After multiple testing correction, we identified 267 genes significantly associated with AsAoD (table S5) including genes well described in mendelian diseases affecting the aorta *ELN, JAG1, EDN1, LOX*, and *FBN1*. We further evaluated the association of imputed tissue-specific gene expression levels with AsAoD risk across multiple tissues and cells using S-PrediXcan. We then used COLOC to formally assess the posterior probability of true colocalization events between gene expression and AsAoD. Predicted levels of fibroblasts growth factor 9 (*FGF9*) which has been shown to play an important role in aorta development, and *PRDM6* levels both associated with AsAoD (figure 2a). We identified 35 gene-tissue pairs (table S7), among which 18 genes colocalize to the expression in the aorta and the strongest were *PRDM6, HMCN1*, and *HAND2* which also displayed expression in coronary arteries, atrial appendage, and left ventricle (figure 2b). Finally, we performed a tissue enrichment association test using MAGMA (implemented in the FUMA (https://fuma.ctglab.nl/)) to identify the set of tissue with significant eQTL enrichment with AsAoD using our GWAS summary statistics and 58 tissues available in Gtex V8. We found significant tissue enrichment for tibial arteries, aorta, coronary arteries, endocervix, colon sigmoid, and ovary which all reflect a predominance of smooth muscle cells in these tissues.

**Figure 2:**
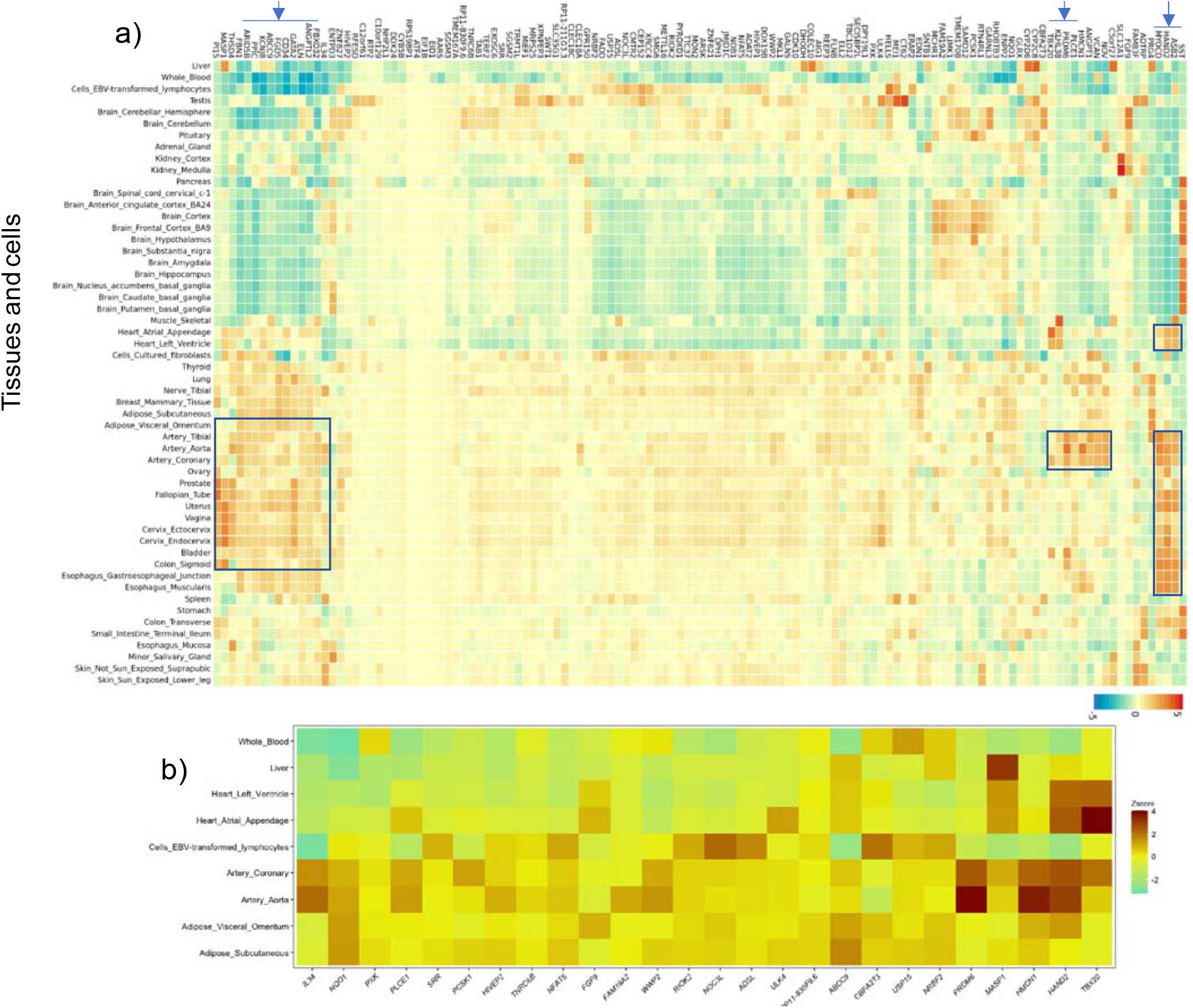
Enrichment of gene-tissue expression pairs within the GWAS of AsAoD (top). The outlines indicate gene sets significantly expressed in aorta and smooth muscle related tissues. (bottom) A subset of gene-tissue pairs significantly enriched in the transcriptome-wide association analysis (S-PrediXcan) and colocalizing in selected tissues of relevance.

### Assessment of pleiotropy

To evaluate the pleiotropic effect of AsAoD risk loci, we extracted all previously reported associations with AsAoD or in high LD with a variant significantly associated with AsAoD from the GWAS catalog. Variants at 31 genomic risk loci were previously associated with multiple phenotypes including aortic root size measured by echocardiography, carotid intima media thickness, abdominal aortic aneurysm, blood pressure (diastolic BP, systolic BP and pulse pressure), coronary artery disease, and a number of blood count measurement and blood biomarkers (figure S5, table S6). We further investigated which of the phenotypes have a high enrichment of GWAS signal in the AsAoD GWAS by comparing the proportion of overlapping genes between the AsAoD and traits reported in the GWAS catalog. We observe a high enrichment of genes previously associated with diastolic blood pressure, pulse pressure, and aortic root size (figure S5), suggesting a high genetic correlation between these traits and AsAoD.

### Genetic correlation

We estimated the genetic correlation between AsAoD GWAS and known clinical risk factors associated with aortic aneurysm such as blood pressure and lipid biomarkers using currently publicly available GWAS summary statistic as well as 238 previously reported traits available on LD Hub v1.9.3 (http://ldsc.broadinstitute.org) (figure S6, table S6). We found a significant genetic correlation between blood pressure and AsAoD (rg=0.31, p.fdr=7.3×10^−19^ for diastolic blood pressure; rg=0.13, p.fdr=45.3×10^−03^ for systolic blood pressure) while no significant genetic correlation was observed for lipid biomarkers. Moreover, among others traits analyzed, we found a significantly high genetic correlation with anthropometric traits including birth weight (rg=0.27, p.fdr=6×10^−11^), height (rg=0.21, p.fdr=4×10^−05^), body mass index (rg=0.18, p.fdr=1×10^−03^); inversely, we found a significant negative correlation between Autism spectrum disorder and AsAoD (rg=-0.27, p.fdr=0.01).

### Polygenic risk score association with Aortic disease

To evaluate whether the enlargement of the aorta can predict the risk of aortic aneurysm, we developed a polygenic risk score (PRS) of AsAoD using the summary statistics of a GWAS performed in a training set of 20,642 unrelated white European using a standard pruning and thresholding method. The PRS was then validated in a separate set of 12,311 UK Biobank participants with cardiac MRI who did not contribute to the original variant effect estimate for the PRS. The association with AsAoD was performed using linear regression adjusting for body surface area (BSA), sex and the top 10 genetic principal components. Sensitivity analyses were performed by stratifying by sex. The PRS with the best predictive performance included 92,299 variants with R^2^<0.8 and p<0.01. Within the validation set a 1 standard deviation (SD) increase in the PRS corresponded to 0.64 mm increase in ascending aortic diameter (β=0.64 (per SD), p=2.5×10^−73^) (figure S7). As expected, the effect estimate was larger in males (β=0.69, p=2.5×10^−38^) than in females (β=0.59, p=2.3×10^−37^) (figure S4).

We examined the association between the PRS of AsAoD and aortic aneurysm in an independent subset of the UK Biobank participants without cardiac MRI measurement at the time of the analysis (n=314,560). Increased PRS for AsAoD was associated with both increased risk of thoracic aortic aneurysm (OR=1.50; p=6.3×10^−03^, figure 3a) and increased risk of surgical intervention (OR= 1.44, p=1.4×10^−03^; figure S7). In a survival analysis, 1 SD increase in the PRS was predictive of 1.42 fold increased risk of thoracic aneurysm (HR=1.42, p=0.01) in the UK biobank (figure 3b). The association with thoracic aortic aneurysm was also replicated in Europeans from the FinGenn biobank, the Million Veterans Project (MVP) and the Penn Medicine Biobank (figure 3a) as well as Hispanic Americans from the MVP, and African Americans from the MVP and Penn Biobanks (figure 3a). The association with PRS AsAoD was even stronger amongst patients who underwent a repair of ascending aortic aneurysm (OR=1.52, p=9×10^−03^; figure S8). Similarly, in the MVP, we observed an OR of 1.57 (p=4.5×10^−03^) among patients of European ancestry, who underwent a thoracic repair after a diagnosis of thoracic aortic aneurysm (figure S8). This finding suggests that, the PRS for larger aortic diameter yields predictive power for thoracic aneurysm that requires eventual surgical intervention.

**Figure 3:**
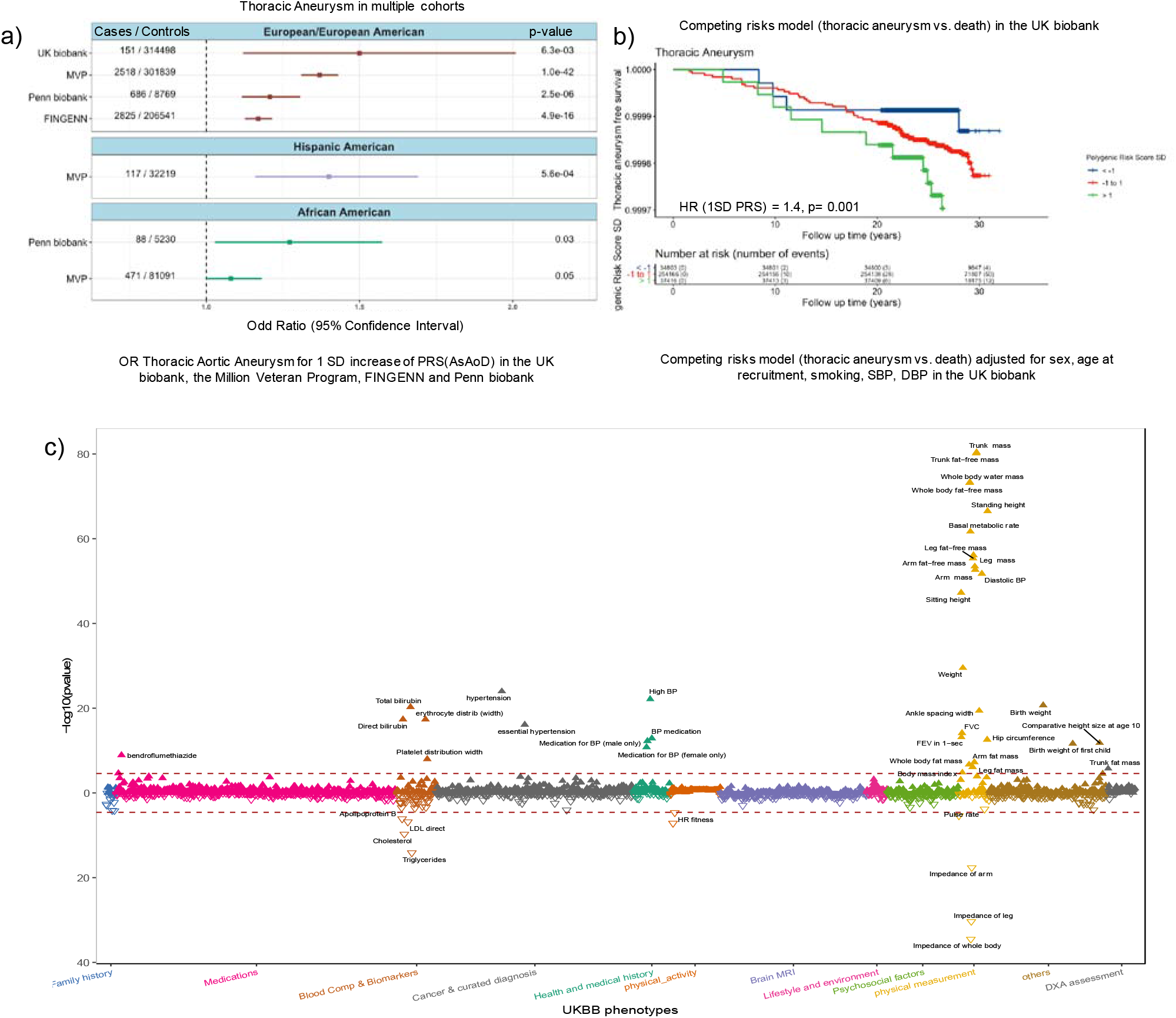
(a) Association between PRS of AsAoD and risk of thoracic Aortic aneurysm and/or thoracic aortic aneurysmal repair across four population-scale biobanks. (b) A competing-risks survival model over a median follow-up time of 29 years for incident thoracic aortic aneurysm (diagnosis or repair) adjusted for smoking, prevalent hypertension, and sex. UKB participants were divided into three tranches based on PRS for display (c) A PheWAS of the PRS of AsAoD and all UKB phenotypes.

We performed a phenome-wide association study (PheWAS) between the PRS of AsAoD on all the UK Biobank phenotypes/traits available in the global biobank engine^18^ and ICD-10 derived phecodes (tables S8 & S9). Overall, increase in physical measurement such as height, weight, body fat percent, and diastolic blood pressure were significantly associated with 1 SD increase in the PRS while lipid biomarkers (low density lipoprotein (LDL), total cholesterol (TC), triglycerides (TG) and Apolipoprotein B) were inversely associated with an increase in the PRS (figure 3). Additionally, an increase in the PRS was associated with an increased risk of hypertension, increase in erythrocyte and platelet counts, and an increase in serum bilirubin levels (figure 3).

### Mendelian randomization

Finally, we investigated the causal relationship between clinical risk factors for ascending aortic dilatation using mendelian randomization (MR) with systematic assessment of heterogeneity and horizontal pleiotropy using MR-presso (tables S10-13, figure S9). Using the inverse-variance weighted method we observed that a genetic 10 mmHg increase in diastolic and systolic blood pressure was causal for a one mm increase in the diameter of the ascending aorta (β=0.01 (p=4.4×10^−06^), and β=0.02 (p=1.6×10^−03^) for both systolic blood pressure (SBP) and diastolic blood pressure (DBP) respectively (figure 4) without showing evidence of heterogeneity or horizontal pleiotropy (table S11). No causal relationship was observed for pulse pressure, LDL, high density lipoprotein (HDL), TC, TG, or lipoprotein(a).

**Figure 4:**
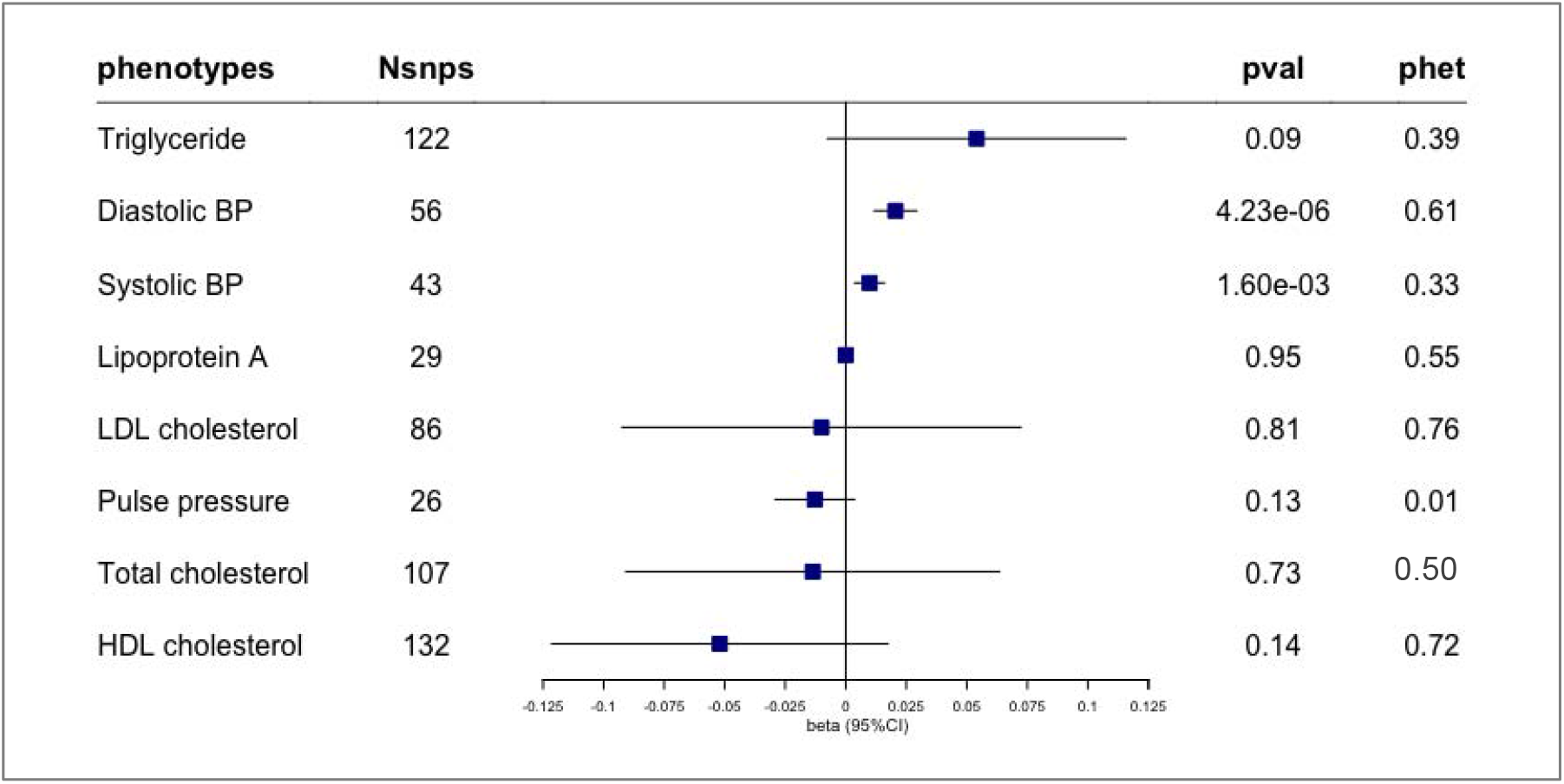
Forest plot depicting causal effect estimates from mendelian randomization analysis testing the association of traditional clinical risk factors with AsAoD. Effect estimates are derived from Inverse Variants Weighted (IVW) analyses and expressed in SD change in AsAoD per SD change in exposure.

## DISCUSSION

In this study, we used a computationally extracted vascular phenotype derived from automated segmentation of prospectively obtained cardiac MRI images from 36,095 UK Biobank participants to characterize the genetic architecture of the diameter of the ascending aorta. The automated measurements of AsAoD in our study population was similar to expected distributions derived from studies of the general population^10,11^. Moreover, our automated measurements scaled anthropometrically as the observed AsAoD was on average higher in males than females and strongly correlated with body surface area ^19,20^.

We identify 99 lead variants across 71 loci genomic loci with across ancestries associated with variation in the ascending aorta diameter among which the strongest signal was found at 7p11.2 near Elastin (*ELN*). *ELN* encodes a structural protein that is a major component of the aortic wall^21,22^ and contributes to the elastic properties of many tissues including skin and lung. Loss of heterozygosity in *ELN* is associated with Williams syndrome characterized by supraventricular aortic stenosis (SVAS) resulting from a loss of elastin formation in the intracellular matrix and consequent weakening of connective tissue and blood vessels^8,23^. Inversely, the 7q11.23 duplication is associated with aortic dilation due to the presence of an additional copy of *ELN* and other genes in the 7q11.23 region^22,24,25^. In addition to *ELN*, several other genes in the locus 7q11.23 such as *CLIP2, GTF2I, GTF2IRD1* and *LIMK1* have also been linked to supraventricular aortic stenosis arising from the 7q11.23 duplication. Chromatin interaction analysis suggested a strong enrichment of regulatory elements in 7q11.23 genomic risk locus which may interact with promotor and/or enhancers of *ELN*. Moreover, we found significant eQTL linking candidate variants within the locus and aortic tissue from GTEx, suggesting the possibility of tissue-specific alterations in gene expression.

Other genes previously associated with aortic diseases among our discovered genomic risk loci include *TBX20* in the locus 7q14 reported in patients with coarctation of the aorta^26^; *PRDM6* (5q23) and *MASP1* (4q34) for which mutation in the genes have been reported in children with patent ductus arteriosus^27,28^. Interestingly, the locus 5q23 (*PRDM6)* along with the locus 17p13 (*SMG6 - SRR*) has been previously related to aortic root size ^6^. Furthermore, our findings highlight multiple independent loci across ancestries in the critical cardiovascular region 4q31-35 harboring *HAND2* a the transcription factor responsible for reprogramming of fibroblast into cardiomyocytes; deletion of this locus have been associated with multiple type congenital heart defect^12-14^. Moreover, targeted deletion of *HAND2* within the neural crest derived cells in mouse models induce defects in the outflow-tract septation and blood vessel formation^29^.

Our findings also highlight loci previously associated with cardiovascular phenotypes including abdominal aortic aneurysm^30^, carotid intima thickness^31^, two independent coronary artery disease loci^32^, and multiple previously reported blood pressure risk loci^33^ (table S3). These findings suggest a shared genetic architecture between AsAoD and cardiovascular diseases and risk factors.

Our gene level association studies identified 147 trait associations among which 19 genes were also found to be significantly enriched in multiple tissues and cell types including heart, blood vessels, and cells transformed fibroblast; highlighting a potential role of these genes in functions related to development, maintenance, or regulation of aorta structure and diameter. This finding is concordant with our tissue enrichment analysis that showed significant enrichment of GWAS signals in the aorta, arteries, heart and other predominantly smooth muscle cell tissues.

AsAoD displayed a high heritability in line with other continuous measurements of human anatomy such as height and head circumference^34^. However, genetic determinants of the aortic outflow tract size are likely under purifying selection as comparative anatomy suggests the physiological demands of cardiac output are relatively invariant across a wide range of mammalian body sizes^35^. In contrast to AsAoD, height demonstrates evidence of different selection pressures within different populations^36^. The genetic architecture underlying measures of high heritability are in concert with our clinical observations for the PRS. The PRS derived from the European GWAS is strongly predictive of disease risk in not only Europeans, but is also reasonably predictive of disease risk in Hispanic and African American populations. As the predictive characteristics of a European-derived PRS in Hispanic and African American populations are likely due in part to the degree of European admixture, the stratified and transethnic meta-analysis offer a unique possibility to improve predictions across populations for a highly heritable trait. While the risk estimates were positive and statistically significant across all replication cohorts, estimates in FinnGen and in the African Americans strata of the MVP appeared smaller which together strongly suggest the need for population-specific studies of AsAoD to derive population-specific risk scores for thoracic aneurysm.

Clinical decision making regarding medical and surgical treatment of thoracic aortic aneurysm centers around the size of the aneurysm and surrounding vasculature^37^. Our data suggest that inheritance of common and rare genetic variation are a primary determinant of risk for aneurysmal dilation. Additionally, the findings from mendelian randomization support the efficacy of blood pressure reduction for primary prevention of thoracic aneurysm^38,39^, and suggest that the reported therapeutic benefits of lipid-lowering therapies on ascending thoracic aortic aneurysms is likely to be confounded^40,41^.

Our study has a number of important limitations. While our analyses of disease risk included multiple diverse populations from the United States, the PRS of ascending aortic diameter was constructed from individuals of European descent. In order to broaden the utility of clinical prediction while simultaneously gaining a more complete understanding of the genetic architecture of cardiovascular anatomy, similar studies be performed across human populations worldwide. While we have identified common variation surrounding known developmental regulators of aortic morphogenesis, our findings also displayed strong overlap the with genetic determinants of blood pressure which may represent physiological exposure of the ascending aorta to a distending force. Of note the UK Biobank enrolled individuals between the ages of 40 to 60 years; a study of aortic diameter performed in infancy might be less reflective of a lifetime of exposure to blood pressure and other aspects of physiology, lifestyle, and growth. Additionally, we strongly suspect that the healthy cohort bias has removed individuals with extremes of aortic diameter or pathology which influences our genetic findings^42^. Measurements of aortic size were automated while are data are consistent with previous population-based samples, our computational approach may inadvertently introduce error. Altogether our findings identify known developmental and physiological connections to aortic biology and pathology and strongly support our conclusion that *bona fide* genetic effects dominate the signals observed.

### Conclusion

Collectively, our data suggest ascending aorta diameter derived from automated measurements is highly heritable with numerous genetic determinants distributed across the entire genome. Many signals are clearly linked to developmental, anthropometric, and physiological characteristics, and a polygenic predictor is strongly predictive of clinically silent and highly morbid thoracic aneurysmal disease across populations. Our data convincingly demonstrate a primary causal role of blood pressure management in reducing the dilation of the aorta and minimizing the risk of progression to aneurysmal disease while strongly suggesting the lack of causal protective effect of lipid lowering therapies. Overall our findings translate the genetic determinants of normal variation in the size of the aorta both forward and backward; simultaneously identifying determinants of early cardiovascular development while also providing a new approach to thoracic aortic disease.

## METHODS

### Design study populations

#### UK Biobank Genetic and Imaging Dataset

The UK-Biobank (UKB) is a publicly available research database combining genomic information with clinical and prospectively obtained imaging data^43,44^. Amongst the 500,000 volunteer participants aged 37-73 years recruited between the years of 2006 and 2010, 100,000 individuals were selected for participation in the imaging study of Brain, Heart, and Abdominal magnetic resonance imaging (MRI). Due to the design of the imaging protocol, (UK Biobank Limited. Information Leaflet: UK Biobank Imaging Assessment Visit http://www.ukbiobank.ac.uk/wp-content/uploads/2017/04/Imaging-Information-Leaflet.pdf) individuals receiving MRI were more healthy than the overall UKB population. The imaging and clinical data are combined with over 90 million Variants, indels and large structural variants was obtained from genotyping and imputation of all 500,000 UKB participants ^45,46^.

#### Ethical statement

Ethical approval for the use of UKB Imaging and clinical data along with consent from participants was obtained by the National Health Service National Research Ethics Service (ref: 11/NW/0382) and data use approved under applications 15860, 13721, and 24983. The MVP, and Penn Biobank were used as external validation of the association between the PRS of AsAoD and thoracic aortic disease.

#### The Million veteran program (MVP)

As previously described^33^ MVP includes participants over age 18 who served in the United States military and are a longitudinal cohort of active users of the Veteran Health Administration (VA) of any age that have been recruited from more than 60 VA Medical Centers nationwide since 2011 with current enrollment at >825,000. Informed consent was obtained from all participants to provide blood for genomic analysis and to access electronic health record (EHR) data within the VA prior to and after enrollment. At the time of our primary analysis, imputed genetic information was available for up to 459,000 participants assigned to white-European; Black-African and Hispanic ancestries using the HARE algorithm^47^

#### The FinnGen Study

A thoracic aortic aneurysm phenotype was derived from the Finnish national hospital registry and death registry using ICD-9 and ICD-10 codes (listed below) as a part of FinnGen project in 176,899 unrelated individuals. Patients and control subjects in FinnGen provided informed consent for biobank research, based on the Finnish Biobank Act. Additional separate research cohorts with study-specific consents collected prior the start of FinnGen (August 2017) were transferred to the Finnish biobanks after approval by Fimea, the National Supervisory Authority for Welfare and Health. Recruitment protocols followed the biobank protocols approved by Fimea. The Coordinating Ethics Committee of the Hospital District of Helsinki and Uusimaa (HUS) approved the FinnGen study protocol Nr HUS/990/2017. The FinnGen project is approved by Finnish Institute for Health and Welfare (THL), approval number THL/2031/6.02.00/2017, amendments THL/1101/5.05.00/2017, THL/341/6.02.00/2018, THL/2222/6.02.00/2018, THL/283/6.02.00/2019), Digital and population data service agency VRK43431/2017-3, VRK/6909/2018-3, the Social Insurance Institution (KELA) KELA 58/522/2017, KELA 131/522/2018, KELA 70/522/2019 and Statistics Finland TK-53-1041-17.

The Biobank Access Decisions for FinnGen samples and data utilized in FinnGen Data Freeze 5 include: THL Biobank BB2017_55, BB2017_111, BB2018_19, BB_2018_34, BB_2018_67, BB2018_71, BB2019_7 Finnish Red Cross Blood Service Biobank 7.12.2017, Helsinki Biobank HUS/359/2017, Auria Biobank AB17-5154, Biobank Borealis of Northern Finland_2017_1013, Biobank of Eastern Finland 1186/2018, Finnish Clinical Biobank Tampere MH0004, Central Finland Biobank 1-2017, and Terveystalo Biobank STB 2018001.

#### The Penn Medicine Biobank

The Penn Medicine Biobank is a genomic and precision medicine cohort of individuals receiving care in the University of Pennsylvania Health System. Currently >60,000 participants have actively consented for linkage of biospecimens with electronic health record data for broad health related research. Participants have been genotyped on either the Illumina QuadOmni platform at the Regeneron Genetics Center, or either the Illumina GSA V1 or GSA V2 platforms at the Children’s Hospital of Philadelphia Center for Applied Genomics. Standard quality control pipelines were employed and the datasets were combined using an imputation based approach^48^ The data was imputed to 1000G Phase3 v5 reference panel using the Michigan Imputation Server^49^ Genetic ancestry was inferred from principal components derived from common, high-quality variants using SMARTPCA. Related individuals were removed from the dataset based on a kinship coefficient of 0.25 or greater.

#### Measurement of ascending aortic diameter in the UK Biobankimaging cohort

At the time of analysis there, the cardiac MRI imaging in the UK Biobank was available for a subset of 36,201 participants released in two batches: at the first imaging visit (started in 2014) where imaging for ~29,000 participants was released, and a second batch where cardiac MRI was release for an additional ~7,201 participants.

The calculation of ascending aorta diameter was obtained from the series labeled *“CINE_segmented_Ao_dist’’* which is a cinematic view of a full cardiac cycle captured on a transverse plane image of the thorax at approximately T4 at the bifurcation of the main pulmonary artery^46^. To calculate the diameter of the ascending aorta we first performed preprocessing step scaling pixel values to fit in the 0-255 range and cropping the images in the series to become a uniform size of 128×128 pixels. We then selected the first image in each series which represented diastolic blood flow. These procedures provided a uniform image size and pixel range to work with when performing calculations. We next applied a Gaussian filter to the image to smooth edges and reduce noise, followed by a second Gaussian filter and a standard Hough circle transform^50^ specifying the desired identification of two circles, the minimum distance between the two circles, and the minimum and maximum radius of the two circles using the open CV package. We used these identified circles as the markers for the measurements of both ascending (circle 1) and descending aorta (circle 2) by calculating the diameter of the circle and multiplying by the image ratio as identified in the DICOM metadata. A publicly available vignette containing related code can be found online (https://github.com/priestlab/aortahoughcircle). After quality control, the ascending aorta diameter (AsAoD) was derived for 36,021 participants (29,000 participants from the first release and 7,021 participants from the second release)

#### Genome wide association study

At the time of analysis 35,062 UK Biobank participants from different ancestry background (including white European, African and Afro-Caribbean, East Asian, and South Asian) had both MRI imaging data and genetic data available. We performed a genome wide association study of the MRI derived AsAoD on unrelated individual within each ancestry group (32,2215 White European, 262 African/Afro-Caribbean, 441 South Asian, and 133 East Asian. We followed by a transethnic meta-analysis of 33031 participants from all ancestry group as well as a meta-analysis on non-White European participant (836 participants). Individual ancestry group where defined using a combination of self-reported ethnicity and genetic information through principal component clustering (figure S10). The association between Variants and ascending aorta diameter were performed using using linear regression in PLINK with adjustment on age, sex, body surface area, and 10 principal components. Variants with MAC lower than 5, imputation quality less than 0.6 and that was deviating from hardy Weinberg equilibrium were excluded from the analysis. As a sensitivity analysis, we also performed the association with AsAoD indexed on the body surface area (BSA) and observed similar results. The transethnic meta-analysis was performed using the classical standard error approach with the software METAL^51^. We applied Bonferroni correction for multiple testing, variants with p<5e-08 were consider significantly associated with AsAoD.

#### Definition of locus and identification independent set of Variants

Genome wide genomic risk locus, lead and candidate variants were identified using FUMA^16^. A genomic risk locus was defined as a 500kb interval in which one or multiple Variants reached genome wide significant threshold. LD blocks within 500kb of each other were merged into one locus. Within each genomic locus, independent variants were defined as those with a correlation less than 0.6 (based on LD structure in 10K white British from the UKB). Finally, candidate variants were defined as those with R^2^>0.6 with an independent variant and a p-value < 0.05. Our set of new genomic risk loci were identified using different step; we first identified genomic risk loci within each ethnic group. Second, we identify the set of genomic risk loci in the transethnic meta-analysis. Third we searched for overlapping between genomic loci from the transethnic meta-analysis and genomic loci identified in each ethnic group. We then categorized our final set of genomic risk loci into 1) replicated across all ethnic group if candidate variants in the locus had consistent effect direction across all group and the heterogeneity p-value > 0.05 in the transethnic meta-analysis; 2) replicated in at least two studies if directionality of the effect were consistent directionality across at least 2 ethnic groups and athe p-value of heterogeneity between 0.05 and 1×10^−06^; and 3) ancestry specific locus if candidate variants in the locus were present in only one ancestry group. Additionally, for a genomic locus that reach genome wide significance at the ancestry specific analysis but was not replicated in the trans-ethnic meta-analysis, we performed a random effect meta-analyses if the directionality of effect was consistent in at least two ethnic groups. Additionally, we performed a step wise procedure implemented in GCTA^52^ to select independently associated Variants followed by a joint conditional analysis to estimate the joint effect the subset of independently associated variant. Standard diagnostics were performed for all association tests (figure S11).

#### Variant functional annotation and chromatin interaction

To further investigate the role of variants with suggestive association, we performed annotation of variants with p-value less than 1e-05 from the discovery cohort using Haploreg version 4^15^. The variant annotation included Enhancer and promoter histone marks, variants located in transcription factor binding sites (TFBS) identified from ChIP-Seq experiments (ENCODE Project Consortium, 2011); potential regulatory motif altered; closest annotated gene; and dbSNP functional annotation. Additional annotation of variants with possible clinical relevance were performed using CLINVAR.

#### Heritability estimate and genetic correlations with other complex traits

We used LD score regression to estimate the heritability explained by the genetic variation captured in our population in the entire genome (global SNP-heritability) as well as the proportion of heritability estimate in different area of the genome (local SNP-heritability)^53^. A genome-wide genetic correlation analysis was performed to investigate a shared genetic basic between Aorta diameter and242 complex traits and diseases. The pairwise genetic coefficients were estimated between our AsAoD summary statistic, and each of the 242 precomputed and publicly available GWAS summary statistics for complex traits and diseases by using LD score regression through LD Hub v1.9.3 (http://ldsc.broadinstitute.org). Additional genetic correlation was computed using LDscore regression for blood pressure phenotype based on the summary statistic of Warren et al^54^ which was not available in the LD Hub v1.9.3 repository. Each pair of genetic correlation were considered significant if the p-value were less 1×10^−04^ based on Bonferroni correction for multiple testing.

#### Gene, pathway, and tissue enrichment

To prioritize genes, pathways and tissues involved in the genetic architecture of Ascending aorta morphology, we conducted a number of downstream analysis which involved several analytical approached but all using the GWAS summary statistics of AsAoD. The Multi-marker Analysis of GenoMic Annotation (MAGMA) v1.09 gene, gene-set, and tissue expression analysis, implemented in FUMA^16,17^ was used to perform gene, gene set, and tissue enrichment analysis. Finally, S-PrediXcan^55^ was used to predict tissue-specific gene expression and association with Aortic tissue with eQTL summary statistics for 52 tissues including 48 tissues from GTEx V8 as reference panel. These analyses incorporated genotype covariance matrices based on a random sample of 10,000 UKB participants to account for LD structure. Colocalization analysis was performed to address the issue of LD-contamination in S-PrediXcan analyses. Input data were identical to those evaluated by S-PrediXcan and colocalization was restricted to only variants included in the gene as designated by S-PrediXcan.

#### Polygenic risk score modeling

We developed a weighted polygenic risk score (PRS) of AsAoD to predict the risk of aortic aneurysms and their risk factors. The PRS was developed based on a summary statistic of a subset of 20,642 UKB White European then tested on the remaining 11,573 unrelated UKB white European with ascending aorta diameter available from the cardiac MRI. The weighted polygenic risk score (PRS) was defined as the sum of the allelic effects (β) of variants contributing to the variation to the ascending aorta diameter across the genome for each individual. The number of variants to include in the polygenic risk scoring was defined using a pruning and thresholding methods implemented in PLINK version 1.90. Briefly, the algorithm (--clump) used a linkage disequilibrium-driven clumping procedure to form “clumps” around variants associated with variation in ascending aorta diameter. We defined a R2 threshold of 0.8 for clumping and a p-value threshold of 0.01. To maximize the PRS replication on external sample, insertions-deletions were removed before the clumping procedure. A final set of 92,299 variants were included in the PRS calculation. For each variant contributing to the PRS, the assigned weight was to the effect size estimate (β) of the risk allele for the corresponding variant derived from the AsAoD summary statistic of the discovery stage. The genomic risk score for each individual was estimated by summing the number of alleles carried by each individual multiplied by the weight (β) as follows:

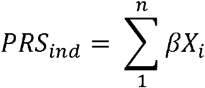

Where PRS_ind_ represents a polygenic risk for a given in individual, n represents the total number of variants included in the PRS, and β is represent the weight (effect size) for the risk allele X of a variant *i*. Xi can take the value 0 (no risk allele), 1 (one risk allele), or 2 (two risk alleles) corresponding to the number of risk alleles X carried by an individual for the given variant. To account for potential residual variability of the PRS in the target population, we centered the score sum of PRS to a mean of 0 and a standard deviation of 1 (μ=0, SD=1).

We tested and validated our PRS in four separate independent samples. First we examined the subset of 11,573 unrelated UKB white Europeans with ascending aorta diameter available from the first and second instance of MRI Imaging release. Although our validation sample was a subset of the UKB participants, this sample was completely independent from the discovery set from which the AsAoD summary statistic was derived. The association between AsAoD and the PRS was performed using linear regression with adjustment on age, sex, BSA, and the top five principal components.

#### Association between PRS for AsAoD and Aortic diseases

The association between Aortic diseases defined by diagnostic codes and the PRS of AsAoD initially was performed on the subset of 316,640 unrelated white British who did not have MRI imaging data available at the time of primary analysis. This was followed by external validation of the PRS in the Million veteran program (MVP); The Penn Biobank; and The FinnGen Biobank.

#### Clinical modeling of thoracic aortic disease

Each replication cohort obtained phenotypic information from administrative or diagnostic coding related to the specific occurrence of thoracic aortic disease. International Classification of Disease 9 and 10 (ICD-9 codes: 441.1, 441.2, 441.01 and ICD-10 codes: I71.01, I71.1, I71.2); Office of Population and Censuses Surveys (OPCS-4 codes: K33.*, L18.2, L19.2, L20.2, L21.2, L22.1, L27.3, L28.3); and Current Procedural Terminology (CPT: 2107426-2107627, 22116452211647, 42740410-42740416, 42740439, 45887730-45890065). For survival modeling of thoracic aortic aneurysm in the UKB individuals with congenital anomalies of the outflow tracts (ICD-10 Q25), syndromic disease involving the aorta (ICD-10 Q87*, Q96, Q93.82), or traumatic injury to the heart or aorta (ICD-10 S25.0) were excluded.

#### PheWAS of Polygenic risk score

We evaluated the correlation between the genetic architecture of the ascending aorta diameter and all phenotypes available in the UKB using a phenome-wide association approach (PheWAS) in which the genetic predictor is the AsAoD PRS and the outcome is the UKB phenotype. Phenotypes were obtained from the Global biobank Engine^56^ which include blood biomarkers adjusted on Statin intake; diseases diagnosis obtained from a combined ICD10 codes, medication, self-reported medical condition, cancer registry, Brain MRI, physical measurement, medication, family history of diseases, cognitive function, and physical activity. Additionally, diseases diagnosis was obtained using ICD10 derived Phecode^57^.

The association between the PRS and each phenotype was performed on 316,640 unrelated white British with without MRI imaging data available at the time of the analysis using linear regression for continuous traits and logistic regression for binary traits with adjustment on age, sex and the top five principal components.

#### Mendelian randomization

To evaluate a causal relation between enlargement of aorta, blood pressure and blood Lipids values, we performed a Mendelian randomization analyses for four lipid phenotypes: LDL, HDL; TG and TC; and three blood pressure phenotypes: SBP, DBP, and PP. The genetic instrument for each phenotype were defined as variants genome-wide significantly (P<5×10-8) associated the exposure and with R^2^< 0.001. All clumping was performed using the TwoSampleMR package using Europeans from 1000G as reference panel. Genetic instruments were obtained from the summary statistics of Warren et al^54^ and Willer et al^58^, were used respectively for blood pressure and lipid phenotypes. We used the Inverse-variance weighted MR methods for the primary analysis^59,60^, with weighted-median MR performed as sensitivity analysis, allowing for up to 50% of the weight of each instrument to be drawn from invalid instruments while controlling type-I error. We performed diagnostics including leave-one-out, single-SNP, funnel-plot, and MR-PRESSO to evaluate for evidence of heterogeneity and horizontal pleiotropy. Additionally, we performed the MR-PRESSO^61^ test to identify evidence of horizontal pleiotropy, test for a global horizontal pleiotropy, and identify outliers followed by MR re-estimate after outlier correction, and finally by performing a test for distortion which consist of testing if the causal estimate is significantly different after outlier adjustment.

## Data Availability

All data from the UK Biobank are publicly available to qualified researchers at the UK Biobank website https://www.ukbiobank.ac.uk/

## Acknowledgements

The authors wish to thank Daniela Zanetti PhD, Matthew Aguirre AB, and Mengyao Yu PhD for technical assistance with aspects of the analysis, and Risto Kajanne for assistance with administrative aspects of the FinnGen resouce.

## REFERENCES

1. Stojanovska, J., Cascade, P. N., Chong, S., Quint, L. E. & Sundaram, B. Embryology and imaging review of aortic arch anomalies. Journal of Thoracic Imaging 27, 73–84 (2012).

2. Lopez, L. et al. Relationship of Echocardiographic Z Scores Adjusted for Body Surface Area to Age, Sex, Race, and Ethnicity: The Pediatric Heart Network Normal Echocardiogram Database. Circ. Cardiovasc. Imaging 10, (2017).

3. Lemaire, S. A. & Russell, L. Epidemiology of thoracic aortic dissection. Nature Reviews Cardiology 8, 103–113 (2011).

4. Aday, A. W., Kreykes, S. E. & Fanola, C. L. Vascular Genetics: Presentations, Testing, and Prognostics. Current Treatment Options in Cardiovascular Medicine 20, (2018).

5. Saeyeldin, A. A. et al. Thoracic aortic aneurysm: unlocking the "silent killer” secrets. General Thoracic and Cardiovascular Surgery 67, (2019).

6. Wild, P. S. et al. Large-scale genome-wide analysis identifies genetic variants associated with cardiac structure and function. J. Clin. Invest. 127, 1798–1812 (2017).

7. McBride, K. L. et al. Inheritance analysis of congenital left ventricular outflow tract obstruction malformations: Segregation, multiplex relative risk, and heritability. Am. J. Med. Genet. 134 A, 180–186 (2005).

8. Pinard, A., Jones, G. T. & Milewicz, D. M. Genetics of Thoracic and Abdominal Aortic Diseases: Aneurysms, Dissections, and Ruptures. Circulation Research 124, 588–606 (2019).

9. Renard, M. et al. Clinical Validity of Genes for Heritable Thoracic Aortic Aneurysm and Dissection. J. Am. Coll. Cardiol. 72, 605–615 (2018).

10. Elefteriades, J. A. et al. Indications and imaging for aortic surgery: Size and other matters. J. Thorac. Cardiovasc. Surg. 149, S10–S13 (2015).

11. Paruchuri, V. et al. Aortic Size Distribution in the General Population: Explaining the Size Paradox in Aortic Dissection. Cardiology 131, 265–272 (2015).

12. Xia, M., Luo, W., Jin, H. & Yang, Z. HAND2-mediated epithelial maintenance and integrity in cardiac outflow tract morphogenesis. Dev. 146, (2019).

13. Strehle, E. M. et al. Genotype-phenotype analysis of 4q deletion syndrome: Proposal of a critical region. Am. J. Med. Genet. Part A 158 A, 2139–2151 (2012).

14. Song, K. et al. Heart repair by reprogramming non-myocytes with cardiac transcription factors. Nature 485, 599–604 (2012).

15. Ward, L. D. & Kellis, M. HaploReg: a resource for exploring chromatin states, conservation, and regulatory motif alterations within sets of genetically linked variants. Nucleic Acids Res. 40, D930–4 (2012).

16. Watanabe, K., Taskesen, E., Van Bochoven, A. & Posthuma, D. Functional mapping and annotation of genetic associations with FUMA. Nat. Commun. 8, 1–11 (2017).

17. Watanabe, K., Umićević Mirkov, M., de Leeuw, C. A., van den Heuvel, M. P. & Posthuma, D. Genetic mapping of cell type specificity for complex traits. Nat. Commun. 10, (2019).

18. McInnes, G. et al. Global Biobank Engine: enabling genotype-phenotype browsing for biobank summary statistics. bioRxiv 304188 (2018). doi:10.1101/304188

19. Davis, A. et al. Diameters of the normal thoracic aorta measured by cardiovascular magnetic resonance imaging; correlation with gender, body surface area and body mass index. J. Cardiovasc. Magn. Reson. 15, 1–3 (2013).

20. Pearce, W. H. et al. Aortic diameter as a function of age, gender, and body surface area. Surgery 114, 691–697 (1993).

21. Curran, M. E. et al. The elastin gene is disrupted by a translocation associated with supravalvular aortic stenosis. Cell 73, 159–168 (1993).

22. Angelov, S. N., Zhu, J., Hu, J. H. & Dichek, D. A. What’s the skinny on elastin deficiency and supravalvular aortic stenosis? Arteriosclerosis, Thrombosis, and Vascular Biology 37, 740–742 (2017).

23. Merla, G., Brunetti-Pierri, N., Piccolo, P., Micale, L. & Loviglio, M. N. Supravalvular Aortic Stenosis. Circ. Cardiovasc. Genet. 5, 692–696 (2012).

24. Earhart, B. A. et al. Phenotype of 7q11.23 duplication: A family clinical series. Am. J. Med. Genet. Part A 173, 114–119 (2017).

25. Morris, C. A. et al. 7q11.23 Duplication syndrome: Physical characteristics and natural history. Am. J. Med. Genet. Part A 167, 2916–2935 (2015).

26. Kirk, E. P. et al. Mutations in crdiac T-box factor gene TBX20 are associated with diverse cardiac pathologies, including defects of septation and valvulogenesis and cardiomyopathy. Am. J. Hum. Genet. 81, 280–291 (2007).

27. Atik, T. et al. Novel MASP1 mutations are associated with an expanded phenotype in 3MC1 syndrome. Orphanet J. Rare Dis. 10, 128 (2015).

28. Sirmaci, A. et al. MASP1 mutations in patients with facial, umbilical, coccygeal, and auditory findings of carnevale, malpuech, OSA, and michels syndromes. Am. J. Hum. Genet. 87, 679–686 (2010).

29. Holler, K. L. et al. Targeted deletion of Hand2 in cardiac neural crest-derived cells influences cardiac gene expression and outflow tract development. Dev. Biol. 341, 291–304 (2010).

30. Bradley, D. T. et al. A variant in LDLR is associated with abdominal aortic aneurysm. Circ. Cardiovasc. Genet. 6, 498–504 (2013).

31. Franceschini, N. et al. GWAS and colocalization analyses implicate carotid intima-media thickness and carotid plaque loci in cardiovascular outcomes. Nat. Commun. 9, (2018).

32. Harst, P. van der & Verweij, N. Identification of 64 Novel Genetic Loci Provides an Expanded View on the Genetic Architecture of Coronary Artery Disease. Circ. Res. 122, 433 (2018).

33. Giri, A. et al. Trans-ethnic association study of blood pressure determinants in over 750,000 individuals. Nat. Genet. 51, 51–62 (2019).

34. Yang, X. L. et al. Three Novel Loci for Infant Head Circumference Identified by a Joint Association Analysis. Front. Genet. 10, (2019).

35. Dewey, F. E., Rosenthal, D., Murphy, D. J., Froelicher, V. F. & Ashley, E. A. Does Size Matter?-Clinical Applications of Scaling Cardiac Size and Function for Body Size. Circulation 117, 2279–2287 (2008).

36. Guo, J. et al. Global genetic differentiation of complex traits shaped by natural selection in humans. Nat. Commun. 9, (2018).

37. Goldfinger, J. Z. et al. Thoracic aortic aneurysm and dissection. Journal of the American College of Cardiology 64, 1725–1739 (2014).

38. Milewicz, D. M., Prakash, S. K. & Ramirez, F. Therapeutics Targeting Drivers of Thoracic Aortic Aneurysms and Acute Aortic Dissections: Insights from Predisposing Genes and Mouse Models. Annu. Rev. Med. 68, 51–67 (2017).

39. Muiño-Mosquera, L. et al. Efficacy of losartan as add-on therapy to prevent aortic growth and ventricular dysfunction in patients with Marfan syndrome: a randomized, double-blind clinical trial. Acta Cardiol. 72, 616–624 (2017).

40. Taylor, A. P. et al. Statin Use and Aneurysm Risk in Patients with Bicuspid Aortic Valve Disease. Clin. Cardiol. 39, 41–47 (2016).

41. Toganel, R., Benedek, T. & Chitu, M. Response to Statin Use and Aneurysm Risk in Patients with Bicuspid Aortic Valve Disease. Clinical Cardiology 39, 307–308 (2016).

42. Aguirre, M., Rivas, M. A. & Priest, J. Phenome-wide Burden of Copy-Number Variation in the UK Biobank. Am. J. Hum. Genet. 105, 373–383 (2019).

43. Collins, R. What makes UK Biobank special? Lancet 379, 1173–1174 (2012).

44. Sudlow, C. et al. UK Biobank: An Open Access Resource for Identifying the Causes of a Wide Range of Complex Diseases of Middle and Old Age. PLOS Med. 12, e1001779 (2015).

45. Bycroft, C. et al. Genome-wide genetic data on ~500,000 UK Biobank participants. *bioRxiv* 166298 (2017). doi:10.1101/166298

46. Petersen, S. E. et al. UK Biobank’s cardiovascular magnetic resonance protocol. J. Cardiovasc. Magn. Reson. 18, (2016).

47. Fang, H. et al. Harmonizing Genetic Ancestry and Self-identified Race/Ethnicity in Genome-wide Association Studies. Am. J. Hum. Genet. 105, 763–772 (2019).

48. Verma, S. S. et al. Imputation and quality control steps for combining multiple genome-wide datasets. Front. Genet. 5, 1–15 (2014).

49. Das, S. et al. Next-generation genotype imputation service and methods. Nat. Genet. 48, 1284–1287 (2016).

50. Biasiolli, L. et al. Automated localization and quality control of the aorta in cine CMR can significantly accelerate processing of the UK Biobank population data. PLoS One 14, (2019).

51. METAL: fast and efficient meta-analysis of genomewide association scans. Available at: https://www.ncbi.nlm.nih.gov/pmc/articles/PMC2922887/. (Accessed: 19th May 2020)

52. Yang, J., Lee, S. H., Goddard, M. E. & Visscher, P. M. GCTA: A tool for genome-wide complex trait analysis. Am. J. Hum. Genet. 88, 76–82 (2011).

53. Shi, H., Kichaev, G. & Pasaniuc, B. Contrasting the Genetic Architecture of 30 Complex Traits from Summary Association Data. Am. J. Hum. Genet. 99, 139–153 (2016).

54. Warren, H. R. et al. Genome-wide association analysis identifies novel blood pressure loci and offers biological insights into cardiovascular risk. Nat. Genet. 49, 403–415 (2017).

55. Barbeira, A. N. et al. Exploring the phenotypic consequences of tissue specific gene expression variation inferred from GWAS summary statistics. Nat. Commun. 9, 1–20 (2018).

56. McInnes, G. et al. Global Biobank Engine: enabling genotype-phenotype browsing for biobank summary statistics. *bioRxiv* (2018).

57. Wu, P. et al. Developing and Evaluating Mappings of ICD-10 and ICD-10-CM codes to Phecodes. *bioRxiv* 462077 (2018). doi:10.1101/462077

58. Willer, C. J. et al. Discovery and refinement of loci associated with lipid levels. Nat. Genet. 45, 1274–1285 (2013).

59. Burgess, S., Butterworth, A. & Thompson, S. G. Mendelian randomization analysis with multiple genetic variants using summarized data. Genet. Epidemiol. 37, 658–665 (2013).

60. Burgess, S., Scott, R. A., Timpson, N. J., Smith, G. D. & Thompson, S. G. Using published data in Mendelian randomization: A blueprint for efficient identification of causal risk factors. Eur. J. Epidemiol. 30, 543–552 (2015).

61. Verbanck, M., Chen, C. Y., Neale, B. & Do, R. Detection of widespread horizontal pleiotropy in causal relationships inferred from Mendelian randomization between complex traits and diseases. Nat. Genet. 50, 693–698 (2018).

